# A multimodal cross-attention pathotranscriptome integration for enhanced survival prediction of oral squamous cell carcinoma

**DOI:** 10.1101/2025.10.31.25339218

**Authors:** Kountay Dwivedi, Amirreza Mahbod, Rupert C. Ecker, Klara Janjić

## Abstract

Oral squamous cell carcinoma (OSCC) accounts for a major part of cancer mortality, with survival outcomes highly dependent on early diagnosis. While many approaches have been proposed for OSCC survival prediction, they often rely on unimodal data, which may be suboptimal. In this study, we introduced a unified cross-attention-based deep learning framework that integrates whole-slide histopathology images (WSIs) and transcriptomic data from OSCC patients for survival prediction. The framework employed an autoencoder for transcriptomic feature extraction and a state-of-the-art pathology foundation model—evaluated across five alternatives—to derive WSI embeddings. These embeddings were subsequently integrated using cross-attention and concatenation within a Cox proportional hazards model. The multimodal approach outperformed nearly all unimodal counterparts, achieving a maximum concordance index of 0.780±0.059 with cross-attention and 0.766±0.050 with concatenation. The results indicate that pathotranscriptomic integration could improve survival prediction for OSCC patients. The implementation is available on GitHub at: https://github.com/kountaydwivedi/multimodal fusion.git.

*This work has been submitted to the IEEE for possible publication. Copyright may be transferred without notice, after which this version may no longer be accessible*.

## 1. Introduction

Oral squamous cell carcinoma (OSCC) attributes to approximately 90% of oral cancers [1]. The 5-year overall survival rate of OSCC declines from 70% - 80% at early stages to approximately 35% - 45% at later stages [2]. Early and accurate diagnostics are therefore essential for prognosis in OSCC cases. Deep learning has transformed the landscape of patient prognosis, automating the analysis of H&E-stained histopathological whole-slide images (WSI) for diagnosis [3]. Particularly, largescale computational pathology models can perform diverse medical image analysis tasks such as segmentation, classification and risk stratification [3-10]. While WSIs portray the spatial description of the tumor, the gene expression-based transcriptomic profile unveils the molecular-level attributes of the disease. As both the modalities are tightly associated [3, 11] pathotranscriptome integration combines both data types, yielding more robust prediction. However, histopathology and transcriptome data are naturally distinct, which poses significant challenges for their integration. A WSI, often being a billion-pixel image, makes direct processing with any conventional deep learning model computationally prohibitive [4]. This issue is mitigated by modeling WSIs using multiple instance learning (MIL) concept [12]. In MIL, the WSI is partitioned into tiles or patches to generate patch-level embeddings, which are utilized for downstream tasks [4]. Contrary to WSI, the transcriptome profile is generally represented as a high-dimensional gene expression vector, often leading to curse of dimensionality. This issue can be alleviated by employing a dimensionality reduction approach [11].

In this study, we present a cross-attention-based pathotranscriptomic integration framework for survival prediction of OSCC patients. We initially employed an autoencoder to derive transcriptomelevel embeddings. Next, we leveraged and compared five state-ofthe-art computational pathology models including CTransPath [6], Prov-GigaPath or GigaPath [7], HibouL [8], Virchow [9] and UNI [10] to extract WSI-level embeddings via attention-based MIL (ABMIL) [13]. These unimodal embeddings were subsequently integrated using cross-attention and concatenation strategies for OSCC survival prediction. This is a novel approach study that utilizes a pathotranscriptomic integration based on pathology foundation model features and dimensionally reduced transcriptomic features via an autoencoder for OSCC survival prediction. We benchmark unimodal and multimodal integration strategies and evaluate the five employed pathology models for extracting WSI-level embeddings suitable for integration.

## 2. Methods

The main workflow of our approach is depicted in Figure 1. In the following, we describe the dataset used and provide a detailed description of the proposed method.

**Figure 1.**
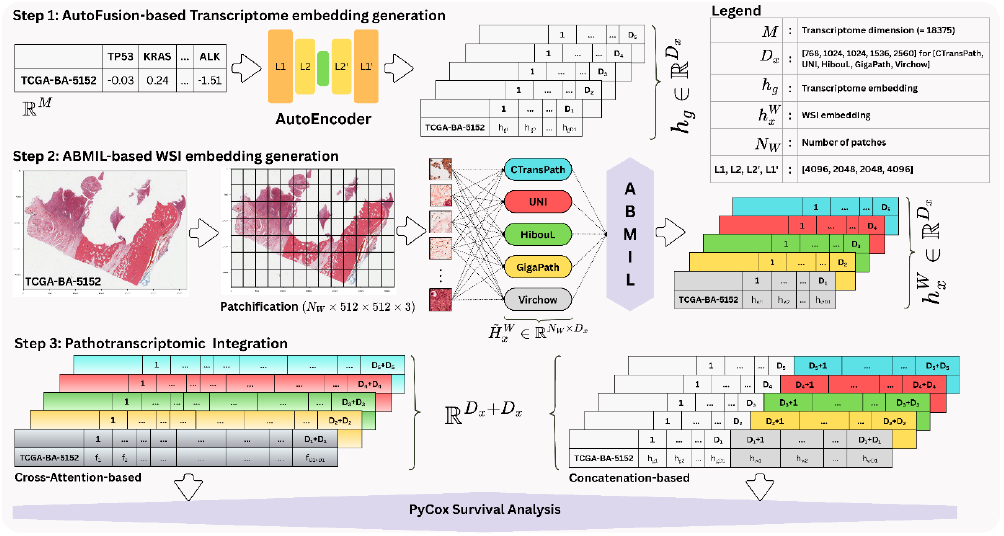
A cross-attention–based deep learning framework for OSCC survival prediction. Step 1: Transcriptome embeddings were derived via autoencoder. Step 2: Patch-level embeddings were computed using distinct pathological models, fusing them to extract WSI-level embedding via attention-based multiple instance learning (ABMIL). Step 3: Transcriptome and WSI embeddings were integrated through concatenation and cross-attention mechanisms to derive pathotranscriptome embedding for subsequent survival analysis.

### 2.1. Dataset acquisition

We utilized the publicly available OSCC dataset provided by The Cancer Genome Atlas (TCGA; https://portal.gdc.cancer.gov/). In total, 522 cases were found under the OSCC subcategory of head & neck squamous cell carcinoma cohort with transcriptome and clinical data (accessed through cBio Cancer Genomics Portal [14]). A subset of n = 38 cases with diagnostic WSIs (DICOM) was selected for experimentation. For clinical attributes, we employed age, gender and the clinical stage of the tumor. The survival outcome was predicted using the overall survival attribute and the performance of the predictive models was assessed by the concordance index (c-index) [15].

### 2.2. AutoFusion-based transcriptome embedding generation

The gene expression transcriptomic data 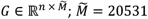, was initially investigated to filter out genes with ≥ 70% missing values across all samples, resulting in a concise *G* ∈ ℝ^*n* × *M*^; *M* = 18375. Next, we z-score normalized *G* across genes to scale its mean and standard deviation to zero and one, respectively.

Given a transcriptome vector *g*_*i*_ ∈ ℝ^*M*^;*i* ∈ [1, *n*], we employed an autoencoder *f*_*a*_ (·) (pretrained on all 522 OSCC cases) to generate a concise embedding vector 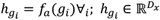. Specifically, a set of four embeddings was derived for *g*_*i*_, each aligned to the dimensionality 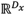 of the respective computational pathology foundation model *x* utilized for WSI embedding generation (see section 2.3.). Subsequently, a transcriptomic matrix 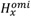 for each *x* was constructed by stacking the respective 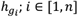 generated by *f*_*a*_ (·) for *x*.

### 2.3. ABMIL-based WSI embedding generation

Initially, each WSI was partitioned into a (512 × 512 × 3) nonoverlapping patches at 40× magnification using the TIAToolbox [16] library. Only patches with tissue area greater than 20% (measured by white– gray pixel thresholding) were selected. The number of patches per WSI varied between 1,510 and 20,393. For a WSI *W*_*i*_; *i* ∈ [1, *n*], assume the set of patches 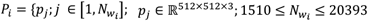. Each *p*_*j*_ was subsequently subjected to each of the five computational pathology foundation models *f*_*x*_(·) to generate its vector embedding map 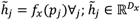 where:

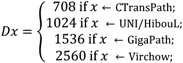

Subsequently, an embedding matrix 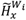 of dimension 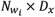 for each *xx* was constructed by stacking all the patch-level embeddings 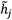 generated by *x*. Thereafter, we employed ABMIL to fuse 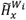 into a single WSI-level embedding vector 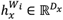 ABMIL initially utilizes a local attention mechanism on the patch-level and subsequently applies a pooling mechanism to generate a WSI-level embedding. For 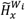, the MIL pooling is computed as [13]:

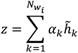

where:

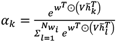

where 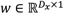 and 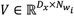 are parameters and ⊙ is *tanh*(·) function.

Finally, a WSI-level matrix 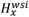 was generated for each *x* by stacking the respective WSI-level embeddings 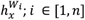 generated by *x*.

### 2.4. Pathotranscriptomic integration

Two strategies were used to fuse the embeddings: cross-attention and concatenation.

#### Cross-attention-based fusion

For each *x* with dimension *D*_*x*_, given vectors 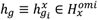 and 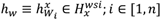, we focused on designing an integration module robust enough to capture the intra-modal and cross-modal correlations. For this, we implemented a modified adaptation of the fusion mechanism proposed in [4] that utilizes the transformer attention [17] as the backbone.

Mathematically, we initialized six vectors: three equal to *h*_*g*_ and three equal to *h*_*w*_ as:

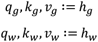

where *q, k* and *ν* denote the *query,key* and *value* vectors, respectively, essentially needed to compute the attention. For the pathotranscriptomic integration, we subsequently computed the vector-based self-attention and cross-attention values and defined a matrix *Attn*_*i*_ as:

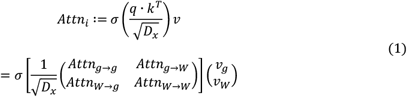

where *σ* is the row-wise softmax. The *q* · *k*^*T*^ captures the intramodality and cross-modality correlations and is mapped as:

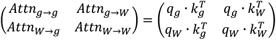

where *Attn*_*g*→*g*_ and *Attn*_*W*→*W*_ represent correlated attentions capturing intra-modal transcriptome-to-transcriptome and histopathology-to-histopathology interactions, respectively, while *Attn*_*g*→*W*_ and *Attn*_*W*→*g*_ represent correlated attentions capturing cross-modal transcriptome-to-histopathology and histopathology-to-transcriptome interactions, respectively. While Jaume *et al*. [4] modeled patch-to-patch interactions using *Attn*_*W*→*W*_ and approximated it with *−∞* in Equation (1) to reduce memory requirements, we explicitly computed this attention expression by utilizing the WSI-level embeddings. The matrix 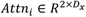 is thereafter normalized using LayerNorm modality-wise and subsequently concatenated to form a final embedding:

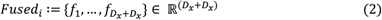

#### Concatenation-based fusion

We extended our methodology by employing a concatenation mechanism to integrate the vectors *h*_*g*_ and *h*_*w*_. For this, we appended both the vectors to form a resultant vector:

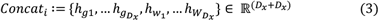

### 2.5. Survival prediction experiments

The computed embeddings *Fused*_*i*_ and *Concat*_*i*_; *i* ∈ [1, *n*] for each *x* are combined with the clinical attributes of *i*. Subsequently, the Cox Proportional Hazard (CPH) model was utilized for survival prediction using the PyCox library [18]. The training was performed using stratified 5-fold cross-validation repeated across 100 different random seed values. Finally, the mean c-index with standard deviation across all seed values was computed for evaluation.

### 2.6. Implementation

#### Autoencoder

The autoencoder utilized in the AutoFusion module comprised two hidden encoder layers of sizes [4096, 2048], a bottleneck layer of size *D*_*x*_ corresponding to the embedding dimension of model *x* and two hidden decoder layers of sizes [2048, 4096], respectively. Each hidden layer was cascaded with a batch-norm layer. We used *tanh*(·) for activation, mean-squared error for loss computation and Adam [19] for gradient optimization. The model was trained for 150 epochs with 1*e*^*−*4^ learning rate and 64 batch size.

#### PyCox

The PyCox-based CPH model was subjected to a vanilla multilayered perceptron (MLP) model with an input layer and a hidden layer of size *D*_*x*_ and an output layer with a single node to output the predictive probability. The MLP model was trained for 500 epochs with early stopping. The optimizer used was AdamW [20] and the batch size was kept to 8.

#### Hardware and Operating System

The entire experimentation was performed on linux operating system installed on a high performance computing GPU server with a dedicated 40GB NVIDIA DGX A100 GPU, 2x AMD EPYC 7742 CPU and 2048GB RWM.

### 2.7. Ablation study

We performed a series of ablation studies to evaluate our proposed framework. For a sample *i* ∈ [1, *n*]:

1. We concatenated full-scale transcriptome vector *g*_*i*_ ∈ ℝ^*M*^ with its corresponding WSI-level embeddings 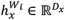,
2. we upscaled the WSI-level embeddings 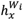 by employing linear interpolation with L2-normalization [21] to match the full-scale transcriptome vector *g*_*i*_ and
3. we concatenated the autofusion-based transcriptome embeddings 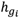 with the WSI-level embeddings generated by two best pathology models: CTransPath 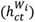 and GigaPath 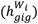

## 3. Results

All benchmark results are provided in Table 1. The unimodal and the multimodal approaches were evaluated on the basis of the mean c-index score (± standard deviation). The per-seed results are provided in the GitHub repository as supplementary materials.

**Table 1.** Evaluation of cross-attention-based pathotranscriptomic fusion against unimodal approaches for OSCC survival prediction. The results are reported based on mean c-index ± standard deviation. Overall, the fusion approach outperformed nearly all unimodal approaches, except GigaPath, where WSI-level embedding exhibited superior results.

| Dimension ($D_x$) | 768 | 1536 | 1024 | 2560 | |
| --- | --- | --- | --- | --- | --- |
| Transcriptome<br>(autoencoder-based embeddings) | 0.692±0.051 | 0.715±0.057 | 0.687±0.051 | 0.736±0.057 |  |
| Model ($x$) | CTransPath | GigaPath | UNI | Hiboul | Virchow |
| WSI | 0.719±0.053 | 0.762±0.059 | 0.715±0.049 | 0.712±0.056 | 0.715±0.060 |
| WSI+Transcriptome<sub>Concat</sub> | 0.766±0.050 | 0.759±0.056 | 0.750±0.055 | 0.737±0.058 | 0.737±0.057 |
| WSI+Transcriptome<sub>Fusion</sub> | **0.780±0.059** | 0.756±0.059 | 0.753±0.054 | 0.756±0.053 | 0.738±0.058 |

Further, the box-and-whisker plots in Figure 2 illustrate the comparison of cross-attention-based and concatenation-based pathotranscriptomic fusion with unimodal approaches.

**Figure 2.**
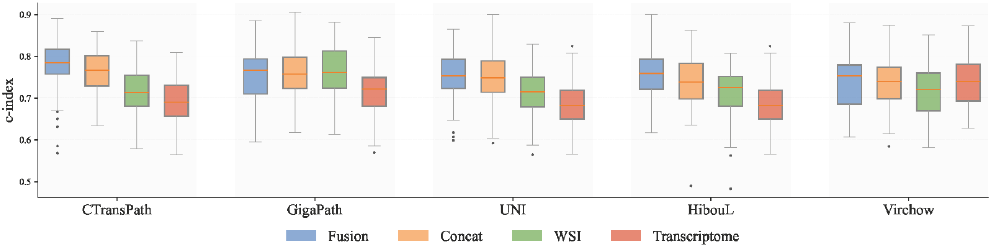
Box-and-whisker plots illustrating the comparison of c-index between cross-attention-based and concatenation-based pathotranscriptomic fusion approaches and unimodal (WSI and transcriptome) models. Except for GigaPath, both fusion mechanisms outperform individual modality-based survival prediction models. Each box-plot summarizes results obtained from 100 independent iterations of the experiment using different seed values.

Overall, both integration strategies outperformed nearly all unimodal approaches. The highest performance was achieved with the cross-attention-based strategy (0.780±0.058), followed by the concatenation-based strategy (0.766±0.050), when combining CTransPath-based WSI features with autoencoder-based transcriptome embeddings. The only exception was the GigaPath model, where WSI-based embeddings exhibited slightly better predictive performance (0.762±0.059) than concatenation-based (0.759±0.056) and attention-based integration (0.756±0.059). The results underscore that histopathology and transcriptome profiles of OSCC patients can be complementarily integrated for better prediction.

Table 2 summarizes the findings of ablation studies. In general, the concatenation of full-scale transcriptome with WSI-level embeddings yielded better results than upscaling WSI-level embeddings and concatenating them with full-scale transcriptome feature set or concatenating transcriptome embeddings with WSI-level embeddings generated by CTransPath and GigaPath. However, none of the ablation-based experiments outperformed the proposed cross-attention-based pathotranscriptomic fusion approach.

**Table 2.** Comparison of different ablation studies with the proposed cross-attention-based fusion approach.

| Ablation | CTransPath | GigaPath | UNI | HibouL | Virchow |
| --- | --- | --- | --- | --- | --- |
| **1. Full-scale transcriptome + WSI** | $0.694\pm0.069$ | $0.714\pm0.063$ | $0.694\pm0.067$ | $0.710\pm0.068$ | $0.667\pm0.067$ |
| **2. Upscaled WSI + full-scale transcriptome** | $0.697\pm0.057$ | $0.689\pm0.054$ | $0.672\pm0.057$ | $0.691\pm0.066$ | $0.681\pm0.065$ |
| **3. Transcriptome + WSI<sub>CTransPath</sub> + WSI<sub>GigaPath</sub>** | | | $0.700\pm0.071$ | | |
| **Proposed (with fusion)** | $0.780\pm0.059$ | $0.756\pm0.059$ | $0.753\pm0.054$ | $0.756\pm0.053$ | $0.738\pm0.058$ |

## 4. Conclusion

This study proposes a novel cross-attention-based pathotranscriptomic integration framework for OSCC profiles, designed to assist pathologists in achieving early and accurate survival prediction. By effectively integrating two complementary modalities - transcriptomic data and WSIs - the proposed framework exhibited superior predictive performance compared to unimodal approaches. These findings confirm that employing a cross-attention-based integration strategy increases the accuracy of survival prediction in OSCC.

## Data Availability

All data produced in the present study are available upon reasonable request to the authors.

https://github.com/kountaydwivedi/multimodal_fusion.git

## Acknowledgements

This work was supported by the Austrian Research Promotion Agency (FFG), project no. 895420.

## Compliance with ethical standards

This study was conducted retrospectively, using publicly available data from the ethically approved TCGA program. Ethical approval for the present study was not required.

